# A Mixed Methods Approach Incorporating Text Analytics to Examine Persistence of Depression

**DOI:** 10.1101/2022.07.19.22276147

**Authors:** Sarah A. Johnson, Andrea Cassells, T.J. Lin, Elisa S. Weiss, Jonathan N. Tobin

## Abstract

**Background:** Some patients’ depression persists despite evidence-based interventions; understanding factors associated with depression persistence could inform screening and treatment. We used a novel mixed-methods approach to examine demographic, clinical, and social factors affecting depression persistence among older, low-income women; we also assessed the utility of this approach for evaluating intervention fidelity.

**Methods:** Data used for this study were generated from a comparative effectiveness study comparing the impact of prevention care management (PCM) versus a collaborative care intervention (CCI) on depression among women overdue for cancer screening: We reviewed 700 care manager logs to identify themes among patients’ experiences and analyzed language use using NVivo^®^’s natural language processing (NLP) functionality. 757 women age 50-64 who screened positive for depression at baseline using the Patient Health Questionnaire (PHQ)-9 and were overdue for ≥1 cancer screening test (breast, cervical, and/or colorectal) participated. All received primary care in XXX Federally Qualified Health Centers. We used NLP to quantify differences in language use across intervention groups and explored how often themes appeared in logs of participants whose depression did not meaningfully improve based on PHQ-9 scores. Differences in demographic, clinical, and social factors were examined.

**Results:** Participants with persistent depression were more likely to discuss pain, fear, and transportation. Asthma and anxiety were associated with lower likelihood of depression remission, while no differences were observed in depression remission rates among those with diabetes or hypertension. Patient-centered words, including “needs” and “feelings”, were more common in the CCI group, while procedure-related words, like “screening” and “mammography”, appeared more frequently in the PCM group.

**Conclusions:** Patient-related factors and social barriers contributed to depression persistence. NLP identified patterns of language use in case logs, suggesting unmet needs among depressed patients. NLP is an efficient, effective method for identifying themes in unstructured text and monitoring intervention fidelity.

## 1. BACKGROUND AND SIGNIFICANCE

Depression is a common condition with serious implications for individual and population health.^1-3^ Depression disproportionately affects women and older adults.^1^ Individuals with depression are more likely to experience poorer health outcomes,^4,5^ engage in health-risk behaviors,^6^ and incur higher healthcare costs.^7^ Depression is also associated with significant societal costs due to decreased worker productivity and absenteeism.^8,9^ The World Health Organization estimates depression and anxiety disorders account for over one trillion dollars in expenditures from productivity losses alone.^10^ Depression among older patients is associated with functional decline,^11^ disability,^12^ increased use of primary care, and reduced health-related quality of life.^7,13^ While effective treatments are available, many patients do not receive them.^14^ Moreover, research suggests that some patients’ depression persists despite evidence-based interventions.^8,15,16^ Improving our understanding of factors associated with depression persistence could inform targeted screening and treatment.

Research suggests factors associated with depression include: personal or family history, exposure to physical or psychological trauma, and chronic medical and psychological conditions such as hypertension, diabetes, asthma, and anxiety.^2,17-19^ Particularly among older adults, social and environmental factors, including isolation, loneliness, and lack of community engagement also play a crucial role in psychological well-being.^17,20,21^ Formal examinations of associations between social determinants of health and psychological well-being are challenging. Social determinants are often not assessed in clinical encounters. When they are, they are rarely captured in structured data fields in electronic health records (EHRs) which limits the utility of traditional quantitative analytic techniques in studying these associations.^22^

Natural language processing (NLP) is an advanced form of qualitative analysis that employs algorithms to interpret and manipulate language. Text analytics is a type of NLP which translates patterns in text into data suitable for thematic and quantitative analysis.^23^ These techniques demonstrate promise in analyzing unstructured clinical data. Orescovik et al. (2017) highlighted the effectiveness of NLP in predicting need for additional services among patients with “high-risk social determinants of health.” ^24^ Edgcomb et al (2021) used NLP to differentiate risk of suicide attempts and self-harm among women with mental illness.^25^

Previously, we completed a randomized controlled comparative effectiveness study among depressed women overdue for at least one cancer screening test (breast, cervical or colorectal) We randomized 757 English or Spanish speaking women to two different interventions. We observed that change in depressive symptoms acted as an effect modifier in the relationship between the intervention and receipt of age-appropriate cancer screening. Those whose depression did not improve or only improved modestly were less likely to be up to date with cancer screening tests at the end of the study.

To better understand why some patients’ depression persisted despite evidence-based interventions, and to ensure that our observations were not the result of contamination between study arms, we used a mixed-methods approach to examine differences in the content of case logs maintained by care managers who delivered the interventions in this study. Our approach incorporated quantitative analysis as well as text analytics. We compared language use between participants in each study arm as well as between participants whose depression had and had not improved significantly at 12-month follow-up. We also explored the impact of selected baseline covariates, including anxiety, chronic medical conditions, and highest attained education level on depression outcomes.

## 2. OBJECTIVES

Our study objectives were to:

1. Identify factors associated with depression persistence by examining differences in the content of case logs maintained by care managers in each intervention arm;
2. Assess fidelity of interventions.

## 3. METHODS

### 3.1 Participant population

Sampling strategy and practice sites are described in detail elsewhere.^27,28^ Our study included 757 women ages 50-64 who screened positive for depression at baseline (PHQ-9 ≥ 8) and were overdue for at least one cancer screening test (breast, cervical, and/or colorectal). All participants received primary care at one of six Federally Qualified Health Centers (FQHCs) in Bronx NY. A total of 379 women were randomized to PCM and 378 to CCI. For this secondary analysis (i.e., focus of this paper), we excluded participants with no PHQ-9 recorded at 12-month follow-up (n=126), and participants without case logs (n=25). There was some overlap between these cases, resulting in 128 excluded participants.

### 3.2 Interventions

Both study arms involved monthly participant intervention calls performed by separate, project-supported care managers who were study staff working on-site at the health centers. The CCI care managers provided support and patient navigation regarding cancer screening needs and mental health, including past and present participation in mental health treatment, including whether the participant was currently taking antidepressant medications and/or participating in therapy. PCM care managers addressed only barriers to cancer screening.

Appendix A contains additional detail about each intervention. To ensure fidelity in the PCM and CCI intervention protocols, separate weekly supervisory meetings were conducted with both PCM and CCI care managers to review the number of calls completed, content of calls, and barriers to implementation.

Care managers for both groups kept electronic narrative records or case logs of each discussion they had with study participants. Data were maintained in secure, password-protected files using REDCAP and included: date, duration, and type of encounter (telephone, in person); screening tests received since last call; upcoming primary care and cancer screening appointments; barriers to cancer screening; and readiness to obtain overdue cancer screening tests.

### 3.3 Data collected

Unstructured data were found in the form of case logs. Structured data were obtained from self-report surveys or the participant’s electronic health record (EHR) and included: PHQ-9,^32^ Generalized Anxiety Disorder (GAD)-7,^33^ age, race/ethnicity, language, education level attained, and diagnoses of asthma, hypertension, diabetes, and hyperlipidemia.

PHQ-9 scores were obtained by study staff at baseline and twelve-month follow-up. Due to variation in participants’ baseline depression status, we sought to detect all clinically meaningful improvements in depression rather than only complete remission of depressive symptoms. Our approach allowed us to capture both modest reductions in depressive symptoms among those with less severe depression at baseline, as well as clinically significant reductions among those with more severe depression at baseline, who in some cases still met criteria for depression according to the PHQ-9. We created two variables corresponding to change in depression symptoms: clinically meaningful improvement (CI) (yes=PHQ-9 ≤ 8 at twelve-month follow-up or a 50% reduction from baseline)^34^ and depression remission (OR) (yes=PHQ-9 <5 at twelve-month follow-up).^35^ Anxiety was defined as a GAD-7 ≥ 10 at baseline.^35^

### 3.4 Unstructured qualitative data analysis

We first conducted a manual review of 700 case logs to identify recurrent themes. Once we reached theme saturation, we created a list of terms mapping to those themes displayed in Table 1. We then conducted textual analysis using NVivo^®^ software for Mac^®^ version 12. We generated word frequencies for the entire sample and examined PCM and CCI groups individually, using targeted queries to compare word frequencies between the two intervention arms to assess fidelity. We used the NVivo^®^ word “stem functionality,” which allows the investigator to search for a group of similar terms with a common stem (i.e., “sick*” which includes: sick, sickness, sickly, sicker, sickest, sickening) and generated word clouds and tables summarizing frequency of word use to examine data within groups. We then used matrices to examine differences in frequency of word use across groups. Word trees were used to facilitate an understanding of how terms were being used and inform our interpretations. We also used NVivo^®^ to create datasets pertaining to the frequency with which themes were mentioned and compared those proportions between participants who experienced/did not experience (a) clinically meaningful improvement; or (b) depression remission.

**Table 1.** Mapping of themes to terms^a^.

| Theme | Select Associated terms for which text was search |
| --- | --- |
| Death | Death, died, passed away, passed on, funeral |
| Depression | Depression, sad, antidepressant |
| Fear | Afraid, fear, scared, terrified |
| Financial issues | Afford, cost, finances, food stamps |
| Housing | Rent, evicted, housing, homeless, apartment, shelter |
| Illness | Sick, exacerbation, surgery, hospitalized, ill, under the weather, health problems |
| Insurance | Insurance, co-pay, coverage, health plan, insurance plan |
| Loneliness | Alone, lonely, isolated |
| Pain | Pain, painful, hurt |
| Substance use | Addiction, substance use, rehab, alcoholic, intoxicated |
| Transportation | Ride, transportation, bus, subway, Access a Ride |

<sup>a</sup>Note: NVivo<sup>®</sup> has the capability to search for all terms with the same stem. This functionality allowed us to conduct our analyses without compiling an exhaustive list of terms.

### 3.5 Quantitative analysis

For quantitative analyses, we used the datasets generated from NVivo^®^ to compute chi squares and compare the frequencies with which specific themes appeared in case logs of those whose depression did versus did not significantly improve and did versus did not remit. We used logistic regressions with depression significantly improved (0=depression not significantly improved, 1=depression significantly improved) and depression remitted (0=depression not remitted, 1=depression remitted) as the outcome variables to examine the impact of age and comorbid chronic conditions, including asthma, anxiety, and hypertension on depression outcomes. We conducted both univariate and multivariate analyses using SAS^®^ version 9.4 for PC.

## 4. RESULTS

### 4.1 Participant characteristics

After excluding participants missing case logs or a PHQ-9 score at 12-month follow-up, there were 629 participants (81%) included in this analysis. The PCM and CCI groups were comparable with respect to age, race/ethnicity, language, and highest level of education attained. Table 2 contains basic descriptive information for these participants. The mean ages were 55.9 years (Standard Deviation (SD)=4.2) and 56.2 years (SD=4.25) in the PCM and CCI groups, respectively. In both groups, for approximately 25% of participants, the highest level of education attained was less than 8 years. 81% of participants in the PCM group and 78% of participants in the CCI group identified as being Hispanic. Spanish was the preferred language for more than half of participants in both groups.

**Table 2.** Demographic characteristics of study participants.

|  | **PCM<sup>a</sup> (n=323)** | **CCI<sup>b</sup> (n=306)** | **p-value<sup>c</sup>** |
| --- | --- | --- | --- |
| **Age mean (SD)** | 55.9 (4.2) | 56.2 (4.25) | 0.1452 |
| **Race/ethnicity** | n (%) | n (%) | 0.4348 |
| Hispanic or Latino | 261 (81) | 238 (78) |  |
| Black/African American | 53(16) | 65(21) |  |
| White | 13 (4) | 12 (4) |  |
| Asian | 1 | 0 |  |
| American Indian/Alaskan Native | 2 (1) | 1 |  |
| **Language** |  |  | 0.7830 |
| English | 131 (41) | 127 (42) |  |
| Spanish | 192 (59) | 178 (58) |  |
| **Education** |  |  | 0.6343 |
| *12 years/completed high school* | 68 (21) | 73 (24) |  |
| *College graduate* | 27 (8) | 23 (8) |  |
| *Less than 8 years* | 70 (22) | 75 (25) |  |
| *Vocational/technical school* | 5 (1) | 7 (2) |  |
| *Postgraduate* | 2 (0.6) | 4 (1) |  |
| *Some college* | 38 (12) | 34 (11) |  |
| *8-11 years* | 110 (34) | 86 (28) |  |

<sup>a</sup>PCM: Prevention Care Management
<sup>b</sup>CCI: Collaborative Care Intervention
<sup>c</sup>p-value: calculated probability

### 4.2 Manual review of case logs

A manual review of case logs revealed a number of different themes. Case-log texts corresponding to each theme are displayed in Table 3.

**Table 3.** Sample case log text and frequency of appearance for salient themes.

| Theme | Sample case log text | Frequency of use |
| --- | --- | --- |
| Illness | *Patient said she is 'fine.' She is still waiting on her surgery.*<br>*Patient stated she did not get to follow through with her appointment for psychotherapy because [her] 4-year- old grandson just had surgery.* | 817 references |
| Financial difficulties | *Patient was sick and experiencing financial difficulties.* | 318 references |
| Substance use | *Patient stated that although it's been hard dealing with her son who is a drug addict, she is doing better.* | 39 references |
| Death | *Her fiancé passed away almost a year ago that she has not fully recovered from and now her sister passed away 2 months ago.* | 100 references |
| Fear | *Patient stated she is "afraid of going to [her] appointments.* | 93 references |
| Transportation difficulties | *The only problem she is having is transportation to and from her appointment.*<br>*Patient stated she "has not come in due to transportation."* | 90 references |
| Housing | *Patient stated she "lives in the basement and is always at the point of getting evicted."* | 125 references |
| Pain | *Patient stated she "is about the same." Her "arthritis hurts every now and then. The pain comes and goes but she's ok."* | 165 references |
| Insurance issues | *Patient has "not attended to mammogram due to insurance losing."* | 258 references |

### 4.3 Differences between study arms and assessment of intervention fidelity

Notable differences were observed between the text from case logs for PCM and CCI arms. The most commonly used terms were: “patient,” “screens,” “cancer,” “up to date,” “barriers,” and “assessment.” For the CCI group, “patient” was the most common term, used over 3,000 times throughout case logs (weighted percentage = 5.6%). In the PCM group, “patient” was the fifth most common term used with a weighted frequency of 2.0%. In the PCM group, “screening” was the most common term with a weighted percentage of 3.7%, whereas “screens” was the second most commonly used term in the CCI group with a weighted percentage of 2.9%. Other notable differences between these groups included higher frequency use of the words, “feelings,” “hospital,” and “needs” among the CCI group, and a higher frequency of the procedure-related words including, “colonoscopy” “mammogram,” “scheduled” among the PCM group. Figure 1 displays word clouds for the case logs of participants in the PCM vs. CCI intervention groups, respectively. The size and position of the word correspond to the frequencies with which each word appears (larger words that are more central are the most frequently used).^1^

**Figure 1.**
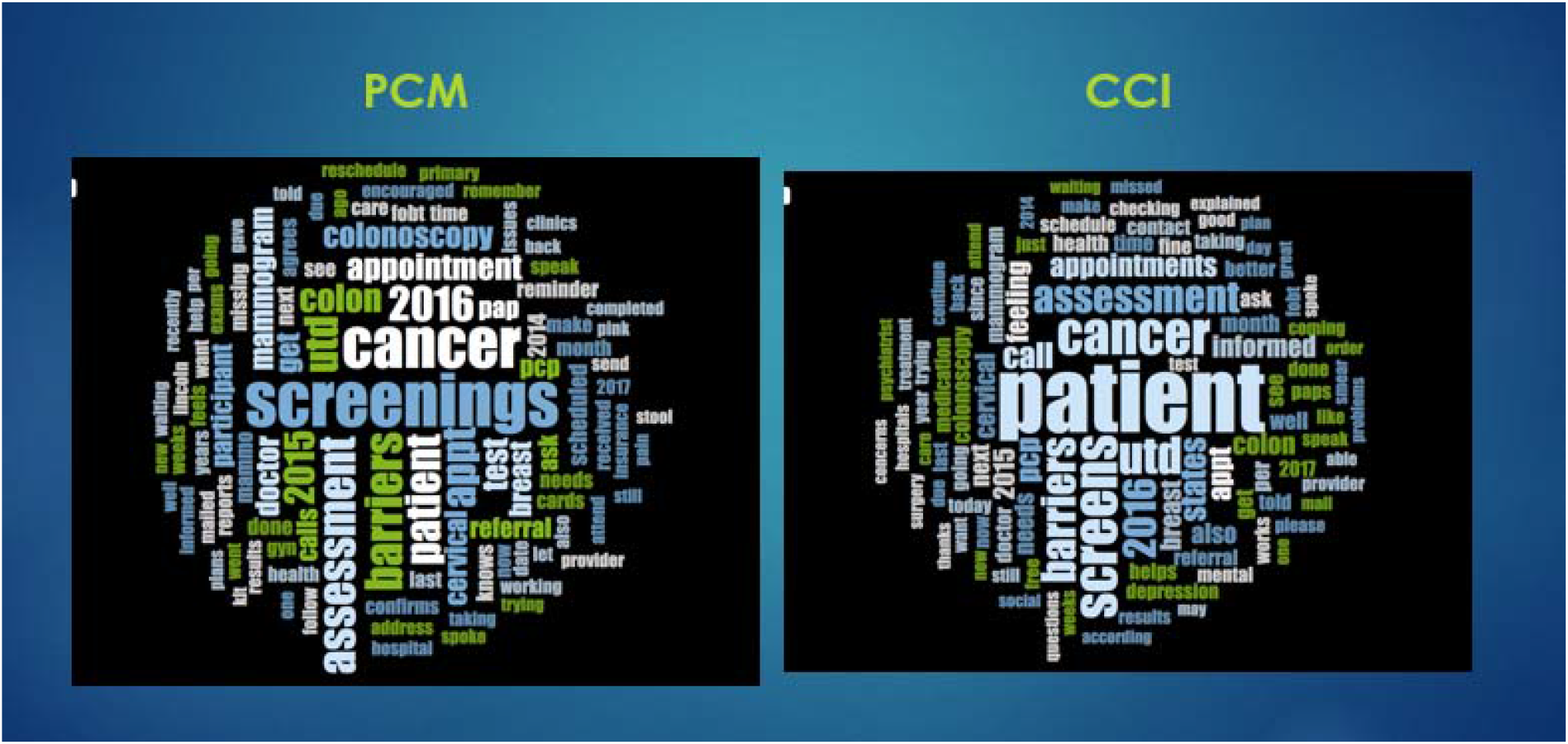
Word clouds representing frequency of word use for PCM vs. CCI study arm case logs. PCM: Prevention Care Management CCI: Collaborative Care Intervention

### 4.4 Differences by baseline PHQ-9 score

The range of PHQ-9 scores at baseline was 8-24 compared to 0-24 at the 12-month follow-up. Nearly 40% of participants (292/763) experienced clinically significant improvements in their depressive symptoms, and nearly 50% (355/763) of participants’ depression remitted; these rates did not differ between intervention groups.

Patients with higher baseline PHQ-9 scores were less likely to experience clinically significant improvements and/or remission of depression symptoms during the study period. The percentage of participants whose depression significantly improved with a baseline PHQ-9 of 9 was 25% lower than that among participants with a baseline PHQ-9 of 8. Similarly, only 4% of participants with a baseline PHQ-9 score of 15 experienced a clinically significant improvement in their depressive symptoms, compared to 16% with a baseline PHQ-9 of 8.

### 4.5 Differences by clinically significant improvement in depression versus not

Figure 2 compares the word clouds for case logs of participants whose depression did and did not significantly improve (because we did not observe a significant difference in depression outcomes between the two intervention arms, we combined both groups for this analysis). On visual inspection, the frequency of word use does not differ between the two groups, with words such as “barriers” appearing with equal frequency.

**Figure 2.**
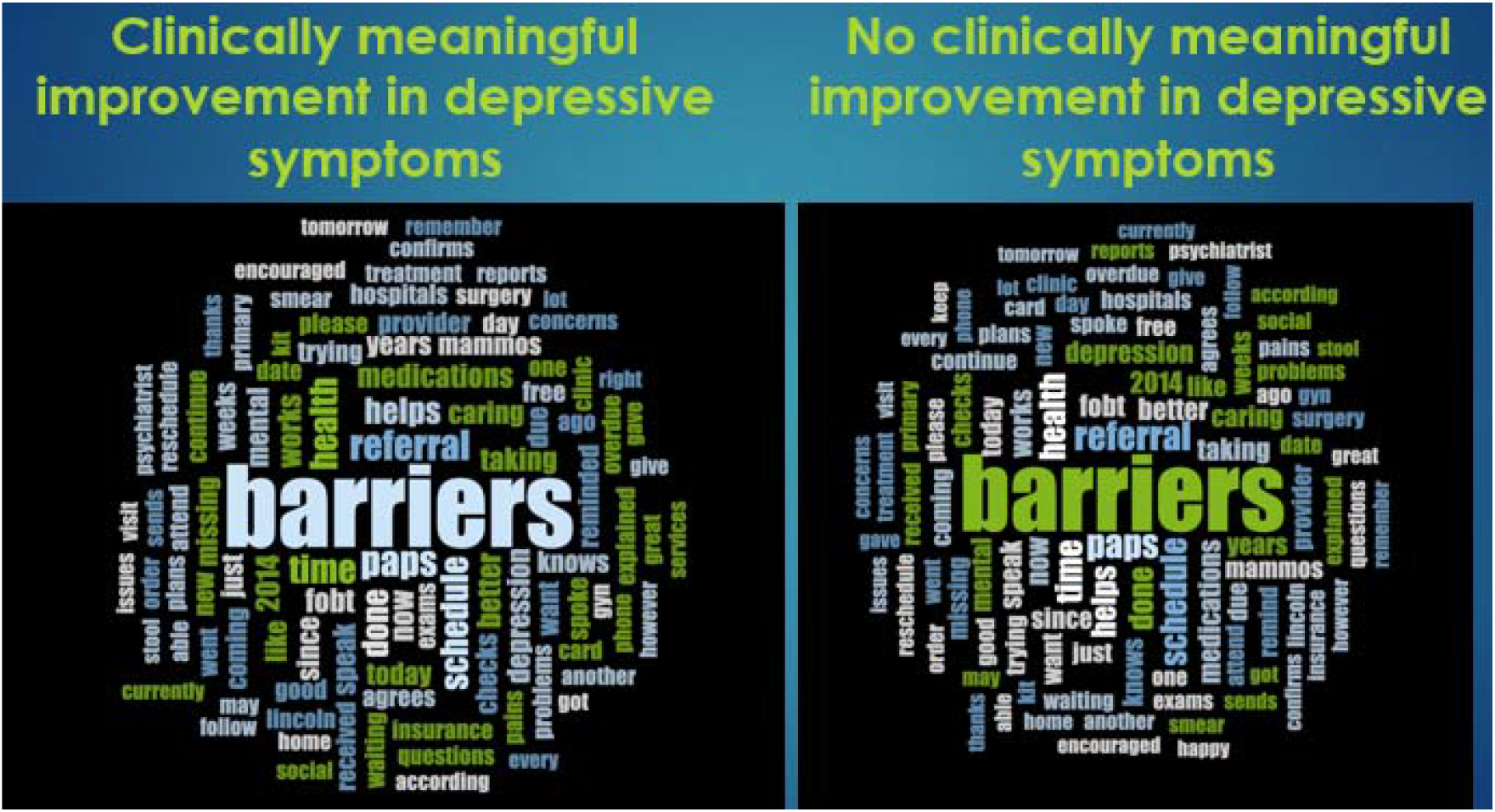
Word clouds representing frequency of word use for patients who did and did not experience clinically meaningful improvement in depression.

Mention of fear was slightly less common among those whose depression improved (OR = 0.83, 95% CI: 0.4, 1.7, chi-square=0.26, *p*=0.61). However, with respect to social determinants of health, those whose depression significantly improved were about a third as likely to report issues related to transportation as those whose depression did not significantly improve (OR=0.3, 95% CI: 0.085, 1.02, chi-square=4.2, *p*=0.04). Those whose depression had improved were nearly twice as likely (OR = 1.7; 95% CI: 1.1, 2.7, chi-square=5.5, *p*=0.02) to discuss financial hardships with their care manager as those whose depression did not.

Table 4 displays differences in the frequency of references to specific themes and number of unique participants referencing those themes among those who did and did not experience a clinically meaningful improvement in their depressive symptoms. At the case-log level, there were differences in the percentages of case logs referencing death, illness, housing, pain, or substance use across these two groups. We observed 176 references to pain among 51 unique patients whose depression had not improve compared to 74 references among 24 unique patients whose depression had improved. There were 459 references to sickness or illness among 123 unique patients whose depression had not improve compared to 358 references among 81 unique patients whose depression had improved.

**Table 4.** Frequency of theme references among unique participants whose depression improved* vs. did not improve.

|  | References among patients<br>whose depression improved<br>(number of patients) | References among patients<br>whose depression<br>had not improved<br>(number of patients) |
| --- | --- | --- |
| Death | 29 (10) | 71 (19) |
| Depression | 198 (42) | 310 (56) |
| Fear | 27 (12) | 66 (23) |
| Financial | 156 (43) | 162 (43) |
| Housing | 62 (24) | 63 (24) |
| Insurance | 135 (35) | 123 (32) |
| Lonely | 4 (2) | 20 (7) |
| Pain | 74 (24) | 176 (51) |
| Sick | 358 (81) | 459 (123) |
| Substance use | 8 (3) | 31 (7) |
| Transportation | 18 (3) | 72 (16) |

### 4.6 Differences between those whose depression remitted vs. not remitted

Participants whose depression had not remitted by the end of study were almost three times more likely to discuss pain (OR = 2.75; 95% CI: 1.4, 5.2) as well as fear (OR = 2.5; 95% CI: 1.0, 6.2). These participants were also more likely to discuss issues related to transportation (OR = 1.6, 95%: CI: 0.61, 4.2). The word “barriers” appeared almost twice as frequently among participants whose depression remitted (3.05% vs 1.87%). References to substance use problems were comparable between groups.

### 4.7 Differences according to comorbidities

Univariate analysis using structured data revealed that participants with comorbid anxiety at baseline were significantly less likely to experience clinically significant improvements in their depressive symptoms at the twelve-month follow-up (OR = 0.34, 95% CI: 0.244, 0.483).

Participants with asthma were about half as likely (OR = 0.58, 95% CI: 0.41, 0.80) as those without asthma to report a clinically significant improvement. Participants with diabetes and hypertension were slightly less likely to experience improvements in their depression symptoms (OR = 0.83 and 0.85, respectively), but neither association was statistically significant. Multivariate analysis revealed that both asthma and anxiety were associated with a significantly lower likelihood of experiencing improvements in depressive symptoms. Participants with asthma were about two thirds as likely (OR = 0.65, 95% CI: 0.458, 0.933) as those without asthma to report remission of depressive symptoms. Neither hypertension nor diabetes appeared to be significantly associated with remission of depression. Multivariate analysis revealed that anxiety was the only factor significantly associated with a decreased likelihood of remission of depression (OR = 0.35, 95% CI: 0.25, 0.50), while asthma approached statistical significance (OR = 0.74, 95% CI: 0.513, 1.076).

## 5. DISCUSSION

In this study of low-income women residing in Bronx NY, multiple patient-related and social factors were associated with improvements and remission of depression over the twelve-month study follow-up period. These included chronic pain, fear, financial issues, and transportation difficulties. Additionally, participants with comorbid anxiety or asthma were significantly less likely to report clinically significant improvements in depressive symptoms or remission of depression at the end of the study. Results from NLP indicated that more frequent use of procedure-focused terms in the PCM arm and patient-centered terms in the CCI arm. This difference provided evidence of the maintenance of intervention fidelity for the two intervention arms. These findings also suggest that NLP can be a valuable tool in analyzing unstructured, qualitative data to assess patient factors as well as intervention fidelity in health services research.

Notably, some differences in language use observed at the case-log level were not statistically significant at the patient level. This suggests certain issues recurred among a small subset of participants whose depression persisted, despite the evidence-based interventions. This finding has important implications for future efforts to leverage the use of NLP-type approaches for analyzing unstructured, qualitative data for identification of high-risk groups and for monitoring treatment outcomes.

The fact that case logs of participants whose depression did not remit were nearly three times more likely to contain mention of pain supports previous work demonstrating a higher prevalence of chronic pain among those with more severe depression.^36^ Tsuji et al (2016) also observed greater functional limitations among participants with comorbid depression and chronic pain, as opposed to those with either condition alone.^37-39^ Similarly, more frequent use of terms related to transportation among those whose depression did not improve or remit adds to previous research which suggests that transportation difficulties can present a major but modifiable barrier to accessing appropriate healthcare services that has a significant impact on clinical, social and emotional well-being, particularly among the older adults.^40,41^ A recent study conducted in the United Kingdom revealed that providing older adults with free bus passes was associated with significant reductions in depressive symptoms and feelings of loneliness.^42^ Many FQHCs provide transportation allowances (subway or bus passes) to enhance access to care and related services^43^; future research should build in coverage for these types of benefits in order to reduce or eliminate access barriers.^44^ This finding also underscores the value of using a community-based approach when designing interventions to address depression, particularly among older patients for whom loneliness and social isolation appear to play a larger role in emotional well-being.^44^

The case logs of those whose depression improved were nearly twice as likely as those whose depression did not improve to mention financial issues. This finding is inconsistent with previous research that found a positive association between financial hardship and depression.^45^ However, this finding within the context of our study may reflect that participants shared information about past financial issues or articulated how they became connected with resources and services that improved their financial situation.

Co-occurrence of both asthma and anxiety was associated with a lower likelihood of both clinically significant improvements in depression and remission of depression. Previous literature supports a higher prevalence of depression among patients with asthma,^46,47^ as well as a higher likelihood of poor asthma control among those with comorbid depression.^48,49^ We are unaware of previous studies, however, reporting that patients with comorbid asthma may also suffer from depression that persists despite intervention. Higher rates of persistence of depression among those with comorbid anxiety could explain high rates of co-occurrence between these conditions throughout the lifespan and argues for the need for better integration of mental and behavioral health services into primary care.^50,51^

We observed clear differences in language use between the CCI and PCM study arms. More frequent use of patient-centered terms such as “patient,” “feelings,” and “needs” in the CCI arm suggest that, as intended, care managers in this study arm were more likely to address issues related to participants’ mental health. In contrast, more frequent use of procedural terms, such as “screening” and “mammogram” indicate that care managers in the PCM arm were more focused on addressing needs related to cancer screening. These findings suggest that efforts to prevent and/or minimize contamination between study arms were successful and lend further support to value offered by NLP, and text analytics in particular, when analyzing unstructured clinical notes. We believe this approach offers an efficient automated approach to demonstrating both clinical process quality and treatment fidelity. Moreover, in the current healthcare climate with increased use of telemedicine, we believe this methodology may be a useful tool in assessing consistency and quality of remote clinical consultations.

This study had several limitations. First, the case logs were manually recorded by the care managers and not by the participants themselves. Due to privacy concerns, calls were not audio recorded. Direct audio recording with automated transcription of the care management calls would have reduced documentation errors. Furthermore, documentation practices may differ between care managers despite their extensive training which might have contributed to selection bias by focusing on known barriers that the intervention was designed to address. Improvements observed in depression over the study period may be due to the natural history of depression or due to regression to the mean, or some combination of both, rather than resulting from the putative mechanisms of either intervention; alternatively, the focus of both interventions on overcoming barriers to accessing care may have had similar non-differential effects on both cancer screening and depression outcomes. Lastly, there may be limitations of using more traditional quantitative analytic approaches when interpreting patterns in language use. More work is needed to examine the utility of these methods in analyzing unstructured, qualitative data. In developing our coding scheme for textual analysis, we relied on our judgment to develop a group of search terms associated with each theme, which may have compromised the sensitivity of our results. Future work could incorporate weighting in order to capture and fully assess the salience of certain themes between groups.

## 6. CONCLUSIONS

This study highlights the importance of certain patient-related and social factors such as chronic pain, comorbid chronic conditions (e.g., anxiety and asthma), and access barriers such as transportation in predicting depression improvement among older, low-income women. Moreover, using text analytics, we identified different patterns of language use between intervention arms, as well as between those whose depression improved or remitted versus those whose depression did not change. This suggests these emerging computerized qualitative methods, when combined with quantitative data in a mixed methods study, can provide novel insights into a variety of methodologic and quality of care issues, including automated monitoring of intervention fidelity and prediction of response to treatment.

## CLINICAL RELEVANCE STATEMENT

This study demonstrates the utility of natural language processing in understanding factors underlying depression persistence in a sample of women ages 50-65. Whereas the use of quantitative analysis only did not offer insight into why some patients’ depression persisted one year after study initiation, using NLP to analyze unstructured clinical data allowed us to identify a number of factors, including social determinants of health, that predicted persistence of depressive symptoms.

## MULTIPLE CHOICE QUESTIONS

1. In this study, natural language processing was used to do all of the following, except
  a. Assess fidelity of intervention arms
  b. Assess depression severity
  c. Identify factors associated with depression persistence
  d. Explore differences in language use between study groups

### The correct answer is B

We did not use natural language processing to assess depression severity. The PHQ-9 was used to assess depression severity at both the outset of the study as well as at the conclusion of the study, 12 months later. Changes in depression severity as measured by the PHQ-9 were the primary outcome of this study. We used natural language processing to understand factors associated with differences in our study outcome. NLP was used to assess fidelity of treatment arms. This was done by comparing language use in the case logs of participants in both study arms, wherein we did observe notable differences. NLP was also used to identify factors associated with depression persistence by exploring differences in language use between those whose depressive symptoms did and did not improve.

2) Which of the following factors did this study demonstrate to be associated with depression persistence?
  a. Age
  b. Location where primary care was received
  c. Chronic pain
  d. Gender

### The correct answer is C

Our analyses revealed that repeated mention of pain in study participants’ case logs was associated with an increased likelihood of persistence of depressive symptoms when compared to participants who did not repeatedly reference experiencing pain. Our analyses did not reveal any associations between age and likelihood of depression persistence, nor did they reveal any differences in the likelihood of depression persistence based on where primary care was received. All study participants were women.

## Data Availability

All data produced in the present study are available upon reasonable request to the authors

## ACKNOWLEDGEMENTS

The authors would like to acknowledge XXX, particularly XXX, for administrative support on this project as well as our FQHC XXXs.

We would also like to thank XXX for funding the main trial (XXX) with additional infrastructure support provided through XXX and our Program Officers, XX and XXX for guidance and advice.

## CONFLICT OF INTEREST

The authors declare that they have no conflicts of interest to report.

### 9.1 Funding

XXX

### 9.2 Author contributions

XXX made substantial contributions to the conceptualization and design of the work. XXX played a substantial role in the acquisition of data by creating and maintaining the REDCAP database and extracting data from it for analysis. XXX conducted qualitative and quantitative analysis and drafted work with substantial contributions and editing from XXX. All authors reviewed and contributed to the final manuscript.

## HUMAN SUBJECTS PROTECTIONS

This secondary analysis did not involve human subjects. The primary research study was reviewed by the XXX Institutional Review Board.

## Appendix A.

Intervention Components for Collaborative Care Intervention and Prevention Care Management

**Appendix A. Intervention Components for Collaborative Care Intervention and Prevention Care Management**
|  | CCI | PCM |
| --- | --- | --- |
| **Cancer Screening Component (PCM)** | Educate and increase cancer screening awareness<br>Provide patient navigation<br>Provide motivational interviewing and support to overcome barriers to cancer screening | Educate and increase cancer screening awareness<br>Provide patient navigation<br>Provide motivational interviewing and support to overcome barriers to cancer screening |
| **Mental Health Component (CCI)** | Provide depression education and motivational support to initiate or continue mental health treatment<br>Be an interface between primary care and mental health providers<br>Provide linkage to social services | None |

*Note.* CCI=Collaborative Care Intervention; PCM=Prevention Care Management.

## Footnotes

1 NVivo^®^ filters out common terms, such as: the, to, from, etc.

## REFERENCES

1. Pratt LA, Brody DJ. Depression in the U.S. household population, 2009-2012. NCHS Data Brief. 2014(172):1–8

2. Pratt LA, Brody DJ. Depression and obesity in the U.S. adult household population, 2005-2010. NCHS Data Brief. 2014(167):1–8

3. Wells KB, Stewart A, Hays RD, et al. The functioning and well-being of depressed patients. Results from the Medical Outcomes Study. JAMA. 1989;262(7):914–919

4. Blumenthal JA, Lett HS, Babyak MA, et al. Depression as a risk factor for mortality after coronary artery bypass surgery. Lancet. 2003;362(9384):604–609

5. Jiang W, Blumenthal JA. Depression and ischemic heart disease: overview of the evidence and treatment implications. Curr Psychiatry Rep. 2003;5(1):47–54

6. Centers for Disease Control and Prevention. Mental Health in the United States: Health Risk Behaviors and Conditions Among Persons with Depression—New Mexico, 2003. MMWR. 2005;54(39):989–991

7. Katon WJ, Lin E, Russo J, Unutzer J. Increased medical costs of a population-based sample of depressed elderly patients. Arch Gen Psychiatry. 2003;60(9):897–903

8. Ivanova JI, Birnbaum HG, Kidolezi Y, Subramanian G, Khan SA, Stensland MD. Direct and indirect costs of employees with treatment-resistant and non-treatment-resistant major depressive disorder. Curr Med Res Opin. 2010;26(10):2475–2484

9. Johnston K, Westerfield W, Momin S, Phillippi R, Naidoo A. The direct and indirect costs of employee depression, anxiety, and emotional disorders--an employer case study. J Occup Environ Med. 2009;51(5):564–577

10. World Health Organization. Mental health in the workplace. Available at: https://www.who.int/team/mental-health-and-substance-use/mental-health-in-the-workplace. Accessed 2017

11. Penninx BW, Guralnik JM, Ferrucci L, Simonsick EM, Deeg DJ, Wallace RB. Depressive symptoms and physical decline in community-dwelling older persons. JAMA. 1998;279(21):1720–1726

12. Verhaak PF, Dekker JH, de Waal MW, van Marwijk HW, Comijs HC. Depression, disability and somatic diseases among elderly. J Affect Disord. 2014;167:187–191

13. Unutzer J, Patrick DL, Diehr P, Simon G, Grembowski D, Katon W. Quality adjusted life years in older adults with depressive symptoms and chronic medical disorders. Int Psychogeriatr. 2000;12(1):15–33

14. Waitzfelder B, Stewart C, Coleman KJ, et al. Treatment initiation for new episodes of depression in primary care settings. J Gen Intern Med. 2018;33(8):1283–1291

15. Fava M. Diagnosis and definition of treatment-resistant depression. Biol Psychiatry. 2003;53(8):649–659

16. Souery D, Papakostas GI, Trivedi MH. Treatment-resistant depression. J Clin Psychiatry. 2006;67 Suppl 6:16–22

17. Cole MG, Dendukuri N. Risk factors for depression among elderly community subjects: a systematic review and meta-analysis. Am J Psychiatry. 2003;160(6):1147–1156

18. Chima CC, Salemi JL, Wang M, Mejia de Grubb MC, Gonzalez SJ, Zoorob RJ. Multimorbidity is associated with increased rates of depression in patients hospitalized with diabetes mellitus in the United States. J Diabetes Complications. 2017;31(11):1571–1579

19. U.S. Department of Health and Human Services, National Insititues of Health, National Institute of Mental Health. Depression. [NIH Publication No. 21-MH-8079]. Available at http://www.nimh.nih.gov/health/topics/depression. Accessed 2018

20. Wagenaar D, Colenda CC, Kreft M, Sawade J, Gardiner J, Poverejan E. Treating depression in nursing homes: practice guidelines in the real world. J Am Osteopath Assoc. 2003;103(10):465–469

21. Singh A, Misra N. Loneliness, depression and sociability in old age. Ind Psychiatry J. 2009;18(1):51–55

22. Adler NE, Stead WW. Patients in context--EHR capture of social and behavioral determinants of health. N Engl J Med. 2015;372(8):698–701

23. Lexalytics. Text Analytics. 2018; https://www.lexalytics.com/technology/text-analytics. Accessed January 30, 2019

24. Oreskovic NM, Maniates J, Weilburg J, Choy G. Optimizing the use of electronic health records to identify high-risk psychosocial determinants of health. JMIR Med Inform. 2017;5(3):e25

25. Edgcomb JB, Thiruvalluru R, Pathak J, Brooks JO, 3rd. Machine Learning to Differentiate Risk of Suicide Attempt and Self-harm After General Medical Hospitalization of Women With Mental Illness. Med Care. 2021;59:S58-S64

26. Wang D, Ogihara M, Gallo C, et al. Automatic classification of communication logs into implementation stages via text analysis. Implement Sci. 2016;11(1):119

27. XXX

28. XXX

29. Dietrich AJ, Tobin JN, Cassells A, et al. Telephone care management to improve cancer screening among low-income women: a randomized, controlled trial. Ann Intern Med. 2006;144(8):563–571

30. Jacob V, Chattopadhyay SK, Sipe TA, et al. Economics of collaborative care for management of depressive disorders: a community guide systematic review. Am J Prev Med. 2012;42(5):539–549

31. Thota AB, Sipe TA, Byard GJ, et al. Collaborative care to improve the management of depressive disorders: a community guide systematic review and meta-analysis. Am J Prev Med. 2012;42(5):525–538

32. Kroenke K, Spitzer RL, Williams JB. The PHQ-9: validity of a brief depression severity measure. J Gen Intern Med. 2001;16(9):606–613

33. Spitzer RL, Kroenke K, Williams JB, Lowe B. A brief measure for assessing generalized anxiety disorder: the GAD-7. Arch Intern Med. 2006;166(10):1092–1097

34. McMillan D, Gilbody S, Richards D. Defining successful treatment outcome in depression using the PHQ-9: a comparison of methods. J Affect Disord. 2010;127(1-3):122–129

35. Pfizer. Patient health questionnaire (PHQ) and GAD7 Instructions. https://www.phqscreeners.com/select-screener. Accessed February 12, 2019

36. Stubbs B, Koyanagi A, Thompson T, et al. The epidemiology of back pain and its relationship with depression, psychosis, anxiety, sleep disturbances, and stress sensitivity: Data from 43 low-and middle-income countries. Gen Hosp Psychiatry. 2016;43:63–70

37. Tsuji T, Matsudaira K, Sato H, Vietri J. The impact of depression among chronic low back pain patients in Japan. BMC Musculoskelet Disord. 2016;17(1):447

38. Vietri J, Otsubo T, Montgomery W, Tsuji T, Harada E. The incremental burden of pain in patients with depression: results of a Japanese survey. BMC Psychiatry. 2015;15:104

39. Vietri J, Otsubo T, Montgomery W, Tsuji T, Harada E. Association between pain severity, depression severity, and use of health care services in Japan: results of a nationwide survey. Neuropsychiatr Dis Treat. 2015;11:675–683

40. Fitzpatrick AL, Powe NR, Cooper LS, Ives DG, Robbins JA. Barriers to health care access among the elderly and who perceives them. Am J Public Health. 2004;94(10):1788–1794

41. Syed ST, Gerber BS, Sharp LK. Traveling towards disease: transportation barriers to health care access. J Community Health. 2013;38(5):976–993

42. Reinhard E, Courtin E, van Lenthe FJ, Avendano M. Public transport policy, social engagement and mental health in older age: a quasi-experimental evaluation of free bus passes in England. J Epidemiol Community Health. 2018;72(5):361–368

43. Weir RC, Emerson HP, Tseng W, et al. Use of enabling services by Asian American, Native Hawaiian, and other Pacific Islander patients at 4 community health centers. Am J Public Health. 2010;100(11):2199–2205

44. Park H. Enabling Services at Health Centers: Eliminating Disparities and Improving Quality October 2006.. Paper presented at: New York Academy of Medicine Enabling Services Roundtable Report; September 29, 2005, New York, NY

45. Brinda EM, Rajkumar AP, Attermann J, Gerdtham UG, Enemark U, Jacob KS. Health, Social, and Economic Variables Associated with Depression Among Older People in Low and Middle Income Countries: World Health Organization Study on Global AGEing and Adult Health. Am J Geriatr Psychiatry. 2016;24(12):1196–1208

46. Di Marco F, Santus P, Centanni S. Anxiety and depression in asthma. Curr Opin Pulm Med. 2011;17(1):39–44

47. Sastre J, Crespo A, Fernandez-Sanchez A, Rial M, Plaza V, investigators of the CSG. Anxiety, depression, and asthma control: changes after standardized treatment. J Allergy Clin Immunol Pract. 2018;6(6):1953–1959

48. Ciprandi G, Schiavetti I, Rindone E, Ricciardolo FL. The impact of anxiety and depression on outpatients with asthma. Ann Allergy Asthma Immunol. 2015;115(5):408–414

49. Ahmedani BK, Peterson EL, Wells KE, Williams LK. Examining the relationship between depression and asthma exacerbations in a prospective follow-up study. Psychosom Med. 2013;75(3):305–310

50. Kessler RC, Berglund P, Demler O, Jin R, Merikangas KR, Walters EE. Lifetime prevalence and age-of-onset distributions of DSM-IV disorders in the National Comorbidity Survey Replication. Arch Gen Psychiatry. 2005;62(6):593–602

51. Kessler RC, Birnbaum HG, Shahly V, et al. Age differences in the prevalence and co-morbidity of DSM-IV major depressive episodes: results from the WHO World Mental Health Survey Initiative. Depress Anxiety. 2010;27(4):351–364

